# Influence of high-energy laser therapy to the patellar tendon on its ligamentous microcirculation: a quasi-experiment

**DOI:** 10.1101/2022.09.27.22280424

**Authors:** Andreas Brandl, Christoph Egner, Ursel Reisser, Christian Lingenfelder, Robert Schleip

## Abstract

Laser therapeutic applications, such as the use of high energy lasers (HILT), are widely used in physical therapy, but basic studies on the mechanisms of action of HILT on tendinous/ligamentous tissue are largely lacking. The aim of this study was to investigate microcirculatory changes of the patellar tendon by HILT. 21 healthy volunteers were treated with HILT. Before and after the intervention, as well as 10 minutes later, the microcirculation was measured by noninvasive laser spectroscopy (O2C device). Tissue temperature was recorded at the measurement time points using thermography. Blood flow increased significantly by 86.38 AU (p < 0.001) after the intervention and by 25.76 AU (p < 0.001) at follow-up. Oxygen saturation increased by 20.14% (p < 0.001) and 13.48%, respectively (p < 0.001), whereas relative hemoglobin decreased by 6.67 AU and 7.90 AU, respectively. Tendon temperature increased by 9.45° and 1.94° Celsius, respectively. Acceleration of blood flow by improving the flow properties of erythrocytes and platelets may have caused the results.

## Introduction

Laser therapy applications are widely used in physical therapy. However, there is a lack of empirical research on fascial structures, their variability and adaptability [1]. Non-invasive therapy devices are becoming increasingly important in the context of musculoskeletal rehabilitation. High-intensity laser therapy (HILT) has been little studied in previous research as an intervention method for knee osteoarthritis [2]. Basic studies on the mechanisms of action of HILT on dense parallel-fibred connective tissues (such as tendons or ligaments) are scarce.

Vascular factors have been identified in previous studies as critical for the development of tendinopathies. Reduced blood flow, especially after activity, represents the greatest risk factor. Studies in the Achilles tendon have shown that structural parameters - such as Achilles tendon thickness, tendon structure as analyzed by ultrasound tendon analysis, or foot posture index – do not significantly contribute to the prediction of Achilles tendinopathy [3], whereas microcirculation in the capillary venous area serves as a significant risk predictor.

Furthermore, microcirculation in tendon tissue plays a prominent role in regeneration after injury. In this context, oxygen, blood viscosity, endothelial functionality and blood vessel permeability are important parameters that emphatically influence wound healing [4].

Hypoxia in tendon tissue promotes the production of proinflammatory cytokines, which can be observed in the early stage of the development of tendinopathy and is therefore discussed as a trigger for such a condition. These hypoxia-related proteins are key mediators of cellular inflammation and apoptosis, processes that cause a shift in collagen matrix synthesis toward an increased proportion of type III collagen (which tends to form fibrils with lower mechanical strength). A shift in matrix structure from type I collagen, which makes up 70% of the dry mass in healthy tendon tissue, to type III collagen reduces the resistance of the tendon to tensile forces, thereby favoring ruptures [5]. Hypoxia further causes local acidosis, corresponding to increased intratendinous lactate accumulation, which further negatively influences pathogenesis [6].

Recent studies indicate that tendinopathies are associated with new vascularization with swelling and pain [7]. However, what seems like a paradox could, according to Järvinen [8], also represent persistent preceding hypoxia. Since tissue regeneration requires an adequate supply of oxygen and nutrients, therefore, neovascularization in tendinopathy can be interpreted as a sign of both persistent hypoxia and a failed attempt to repair the tendon. It is known from the fields of cancer research and retinopathy that hypoxia-induced neovessels are hyper-permeable, resulting in vascular leakage and reduced blood flow. This provides insufficient oxygen and nutrients needed for tissue maintenance and possible regeneration [8].

HILT decreases erythrocyte deformability and platelet aggregation, resulting in membrane revitalization, viscosity reduction, and erythrocyte stress adaptation. In addition, the lifespan of erythrocytes and platelets is prolonged and their flow properties are improved under HILT [9–11]. Therefore, HILT is also discussed as a promising method for the treatment of various blood diseases [9–13].

The aim of this study was to investigate microcirculatory changes of the patellar tendon by HILT.

## Methods

### Study design overview

The study was a quasi-experiment with one intervention group. Measurements were taken before and after the intervention and at a 10-minute follow-up according to the SPIRIT guidelines [14]. The study protocol was prospectively registered with the German Clinical Trials Register (DRKS00028155) on 18.02.2022. The study has been reviewed and approved by the ethical committee of the DIPLOMA Hochschule, Germany (Nr. 1021/2021), has been carried out in accordance with the declaration of Helsinki and has obtained written informed consent from the participants [15].

### Setting and participants

The study was conducted in a school of physiotherapists, in a medium-sized city in middle Germany. The number of participants was calculated based on the results of a previous study and set at 18 [16,17]. The acquisition was carried out via direct contact, a notice board, and the distribution of information material in the school.

Inclusion criteria were: (a) A generally healthy constitution; (b) the persons to be treated must have intact thermal sensitivity and be able to perceive and communicate pain; (c) a BMI between 18 and 29.9; (d) female or male subjects aged 18 to 60 years; (e) sitting position for 15 minutes must be pain-free for the subjects.

Exclusion criteria were: (a) generally valid contraindications to physiotherapeutic and osteopathic treatments of the patellar tendon (i.e., fractures, tumors, infections, severe cardiovascular and metabolic diseases); (b) pregnancy; (c) rheumatic diseases; (d) taking medication that affects blood circulation, pain or mind; (e) taking muscle relaxants; (f) skin changes (e.g. neurodermatitis, psoriasis, urticaria, decubitus ulcers, hematoma); (g) surgery or other scars in the patellar tendon region; (h) previous mental illness; (i) surgery in the last three months; (j) prosthetics or internal knee arthroplasty; (k) acute inflammation.

### Study flow

First, the anthropometric data, age, gender, height, and weight were collected by the investigators (AB, RS). See the flowchart in Fig. 1 for the course of the study.

**Fig. 1.**
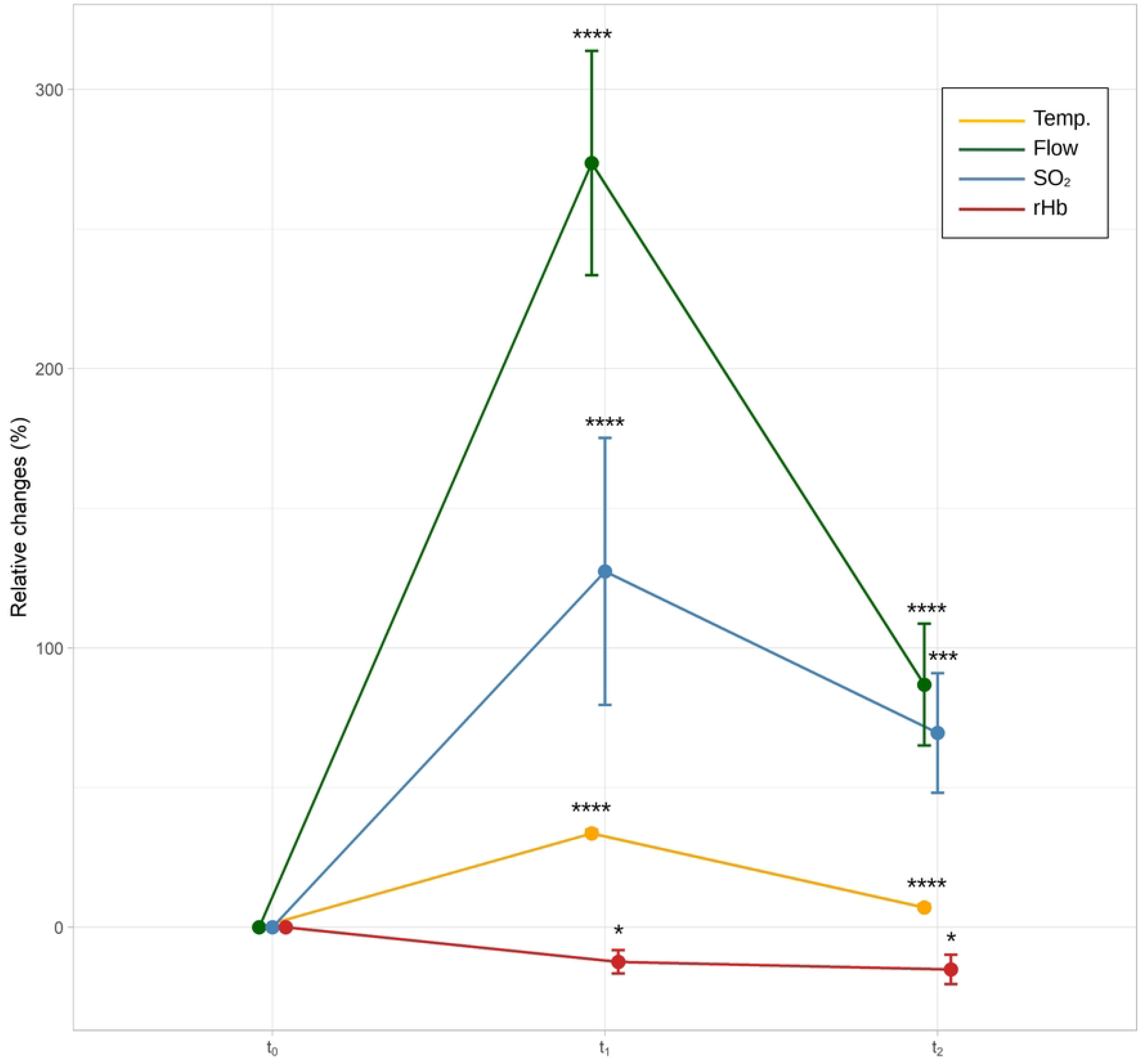
Flow diagram of the study. The “Loss to follow-up” has been removed, as it is not relevant for the study design.

Before the measurements, the subjects were given information regarding the implementation of the intervention and measurements. They lay on their backs with their heads bent at 160° (cervical spine flexion) on a treatment couch. The knees were placed underneath and a knee flexion of 145° - 150° was preset. The measurements were taken in daylight between 9:00 a.m. and 03:00 p.m. and at a room temperature of 22 degrees Celsius. The initial measurement (baseline) was performed first. Then the HILT intervention (OptonPro, Zimmer Medizinsysteme GmbH, Neu-Ulm, Germany, Fig. 2B) was carried out (Fig. 2C). A total amount of energy of 800 J (100 J/cm2) was applied to a predefined area of 2 cm x 4 cm on the patellar tendon (Fig. 2A).

**Fig 2.**
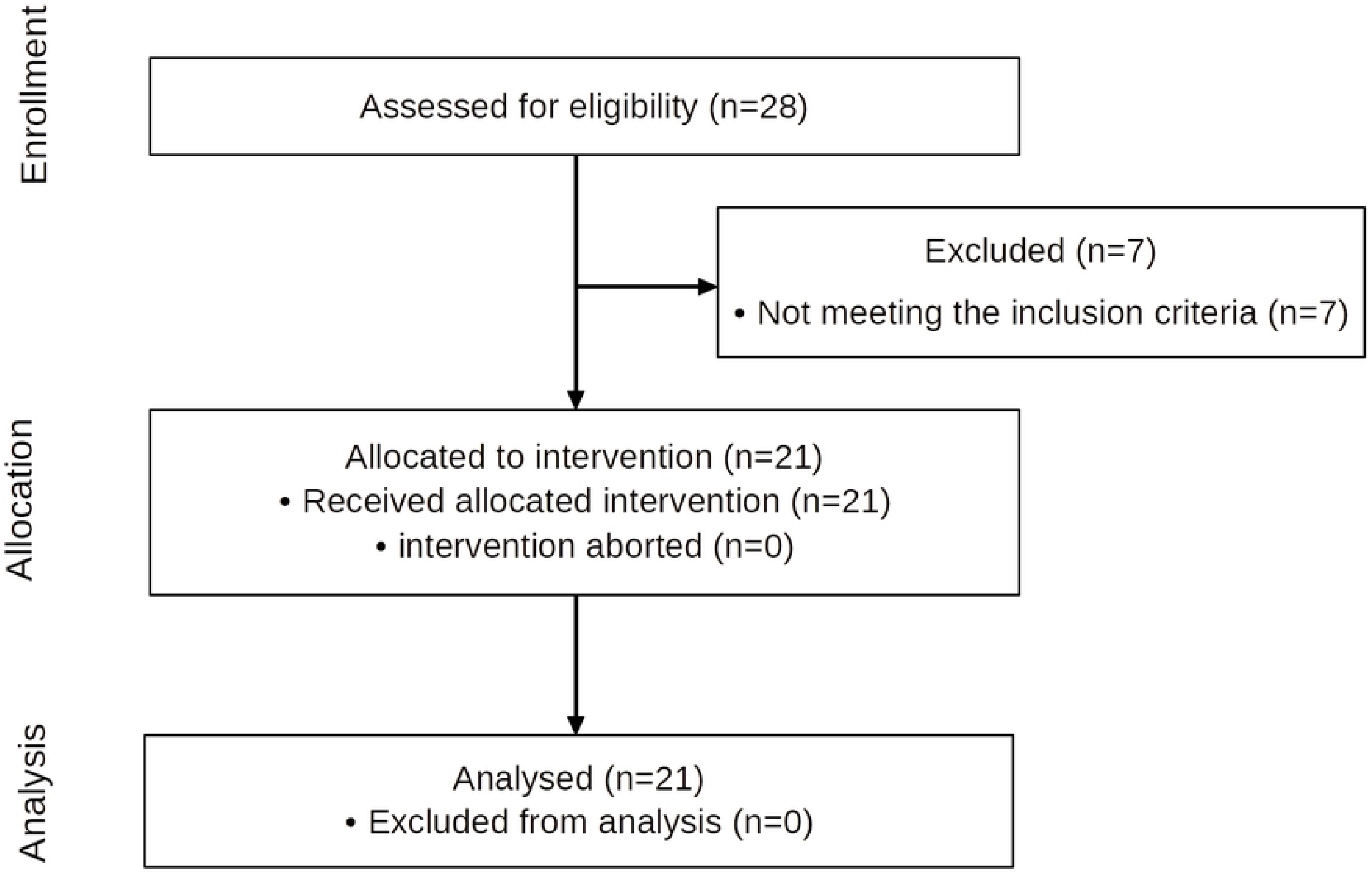
A show the prefabricated template defining the treatment area of 2 cm x 4 cm. B shows the OptonPro high intensity laser therapy device. C shows the laser application to the patellar tendon.

### Outcomes

Measurement of the microcirculation of the patellar tendon was performed using a laser Doppler flowmeter and tissue spectrometer (O2C, LEA Medizintechnik GmbH, Heuchelheim, Germany) (Fig. 3). This makes it possible to simultaneously determine blood flow, oxygen saturation, and relative hemoglobin levels of the measured tissue. The device determines these parameters at the venous end of the capillary and thus provides information about the local microcirculation.

**Fig 3.**
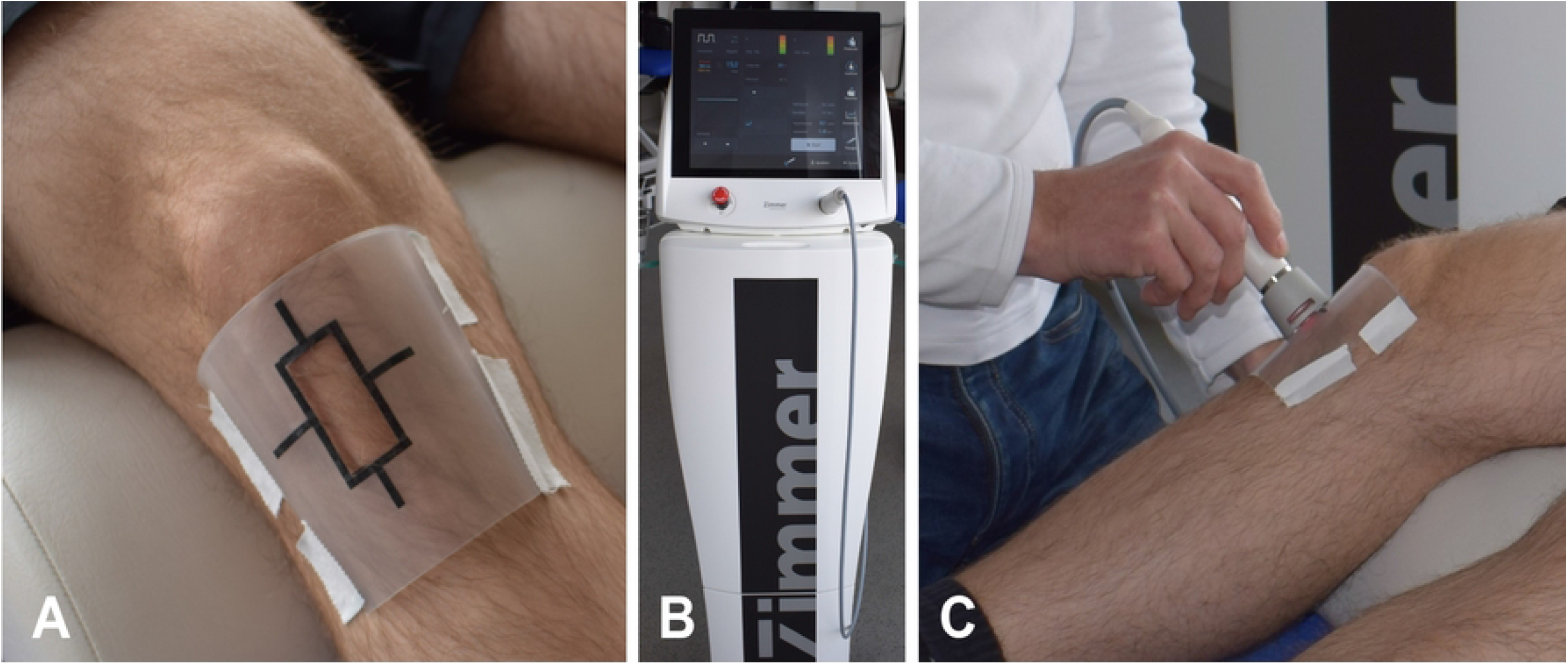
A: Hardware for microcirculation measurement using a laser Doppler flowmeter and a tis-sue spectrometer. B: Probe placement.

Temperature measurement using thermography covers temperature ranges from -20° to 400° Celsius (Flir One, Teledyne FLIR LLC, Wilsonville, US). Measurement accuracy is +/- 3° or +/- 5%. This is valid 60 seconds after switching on the instrument in an ambient temperature of 15° to 35° Celsius and the target temperature is in the range of 5° and 120° Celsius. To ensure measurement at a reproducible measuring point, the template (Fig. 2A) was provided with a marker that ensured exact positioning of the image section for temperature determination (Fig. 4).

**Fig 4.**
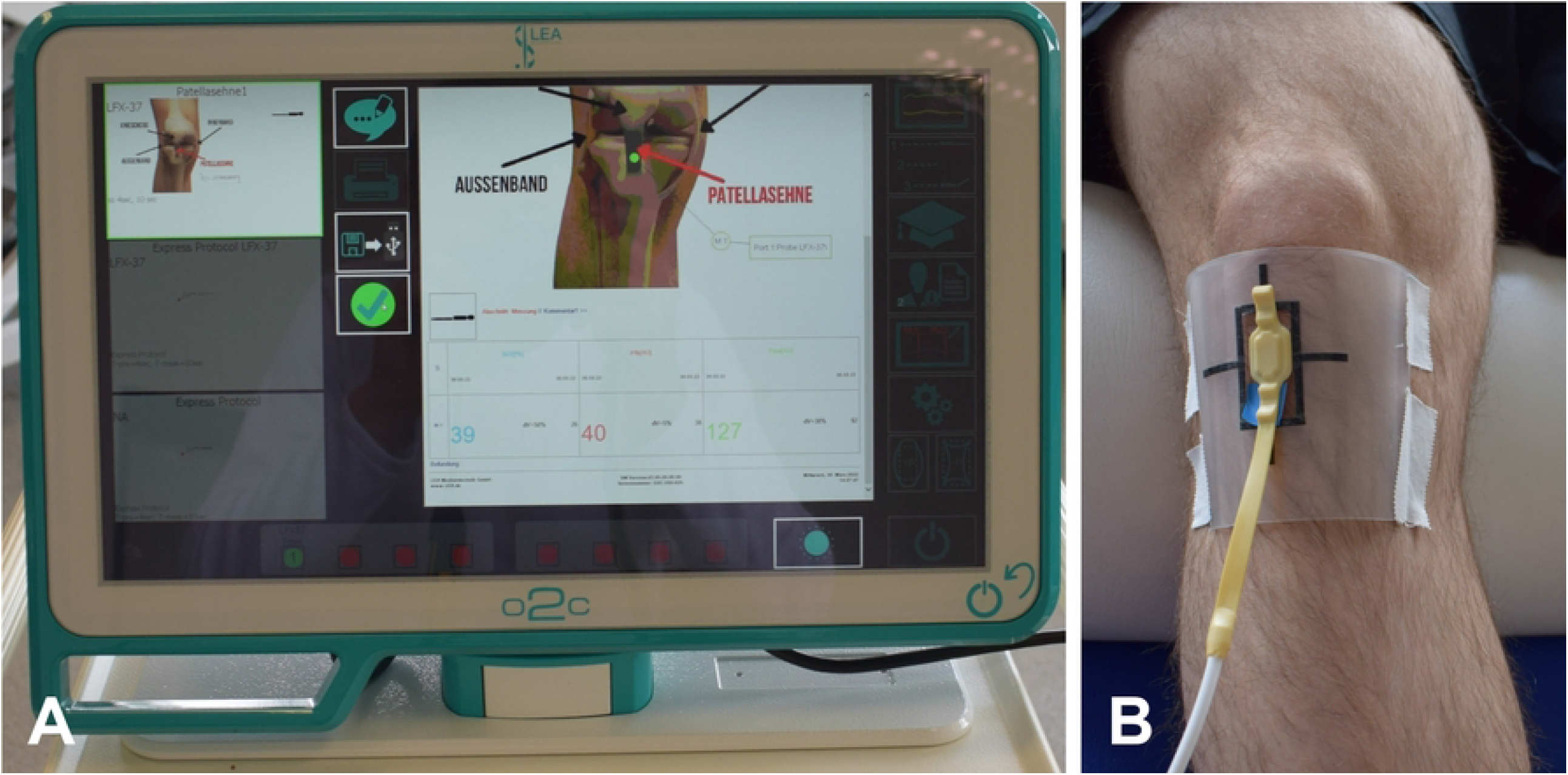
Thermography of the patellar tendon.

### Statistics

The standard deviation (SD), mean, and 95% confidence interval (95% CI) were determined for all parameters. There were no outliers in the data. The outcome variables were normally distributed as determined by the Shapiro-Wilk test (p > 0.05). Differences between measurements were tested for significance using a repeated measures ANOVA. For significant main effects, pairwise comparisons were performed with Bonferroni correction. The significance level was set at p = 0.05.

Libreoffice Calc version 6.4.7.2 (Mozilla Public License v2.0) was used for the descriptive statistics. The inferential statistics were carried out with the software R, version 3.4.1 (R Foundation for Statistical Computing, Vienna, Austria).

## Results

The anthropometric data and baseline characteristics are shown in Table 1. Of 28 subjects screened between 18/02/2022 and 30/03/2022, 21 met eligibility criteria (Fig. 1). None of the subjects reported any adverse side effects of HILT treatment. The raw data on which the study results are based are in the S1 Data Supporting information.

**Table 1.** Baseline characteristics.

| Baseline characteristics | Participants (n=21)<br>mean ± SD |
| --- | --- |
| Gender (men/woman) | 15/6 |
| Age (years) | 22.1 ± 4.2 |
| Height(m) | 1.75 ± 0.1 |
| Weight (kg) | 76.0 ± 11.5 |
| BMI (kg/m<sup>2</sup>) | 24.9 ± 3.5 |
| Sport activity (hours per week) | 6.0 ± 4.2 |

SD standard deviation. n number.

A repeated-measures ANOVA revealed that temperature had a statistically significant difference between measurements, F(2, 40) = 342.46, p < 0.001, partial η^2^ = 0.96. A Bonferroni-adjusted post-hoc analysis revealed significantly (p < 0.001) higher temperature after the intervention (M_Diff_ = 9.45, 95%-CI[8.73, 10.16]) and 10 minutes later (M_Diff_ = 1.94, 95%-CI[1.24, 2.65]) compared to pre-intervention.

There was also a significant difference for blood flow, F(1.56, 31.23) = 116.15, p < 0.001, partial η^2^ = 0.85. A Bonferroni-adjusted post-hoc analysis showed significantly (p < 0.001) higher flow after intervention (M_Diff_ = 86.38, 95%-CI[71.62, 101.14]) and 10 minutes later (M_Diff_ = 25.76, 95%-CI[16.65, 34.87]) compared to before the intervention. These values also exceeded the minimum detectable changes of 9.2 arbitrary units (AU) [3].

A significant difference was found for oxygen saturation (SO_2_), F(2, 40) = 25.01, p < 0.001, partial η^2^ = 0.56. A Bonferroni-adjusted post-hoc analysis revealed significantly (p < 0.01) higher values after the intervention (M_Diff_ = 20.14, 95%-CI[13.41, 26.88]) and 10 minutes later (M_Diff_ = 13.48, 95%-CI[8.10, 18.85]) compared to before the intervention.

A significant difference was found for relative hemoglobin concentration (rHb), F(2, 40) = 7.24, p = 0.002, partial η^2^ = 0.27. A Bonferroni-adjusted post-hoc analysis revealed significantly (p < 0.01) lower rHb values after the intervention (M_Diff_ = -6.67, 95%-CI[-11.46, -1.88]) and 10 minutes later (M_Diff_ = -7.90, 95%-CI[-12.95, -2.86]) compared with the preintervention period.

The changes between the baseline measurement, the measurement after treatment, and the measurement 10 minutes later are shown in Table 2 and Fig. 5.

**Table 2.**
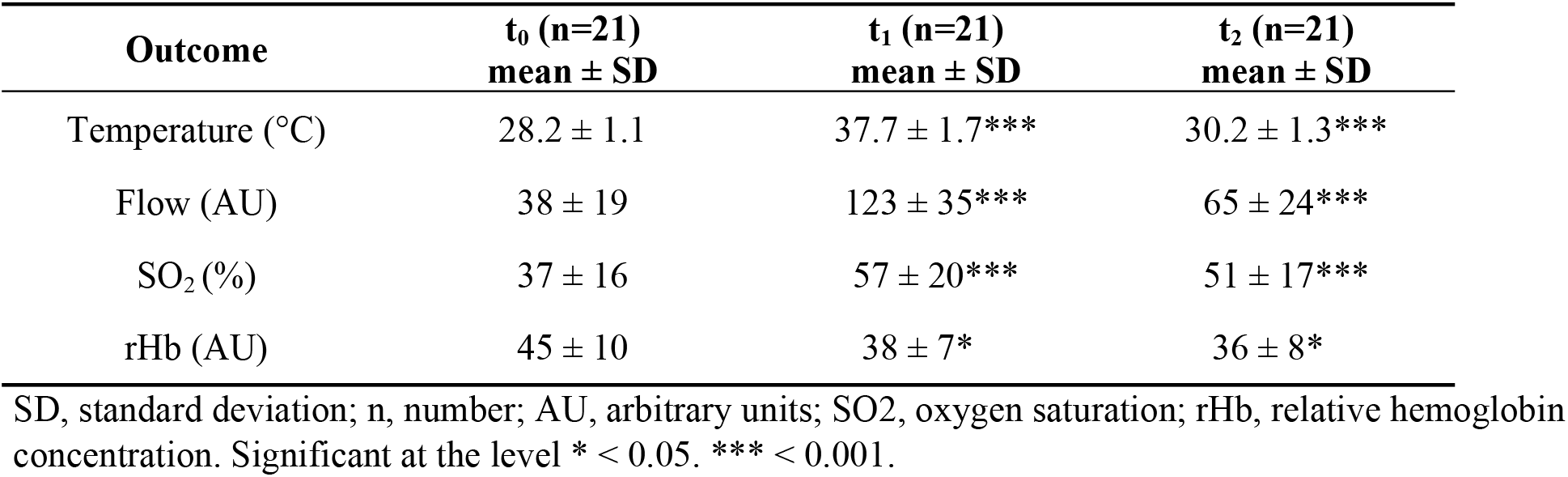
Descriptive statistics.

| Outcome | t<sub>0</sub> (n=21)<br>mean ± SD | t<sub>1</sub> (n=21)<br>mean ± SD | t<sub>2</sub> (n=21)<br>mean ± SD |
| --- | --- | --- | --- |
| Temperature (°C) | 28.2 ± 1.1 | 37.7 ± 1.7*** | 30.2 ± 1.3*** |
| Flow (AU) | 38 ± 19 | 123 ± 35*** | 65 ± 24*** |
| SO<sub>2</sub> (%) | 37 ± 16 | 57 ± 20*** | 51 ± 17*** |
| rHb (AU) | 45 ± 10 | 38 ± 7* | 36 ± 8* |

SD, standard deviation; n, number; AU, arbitrary units; SO<sub>2</sub>, oxygen saturation; rHb, relative hemoglobin concentration. Significant at the level \* < 0.05. \*\*\* < 0.001.

**Fig. 5.**
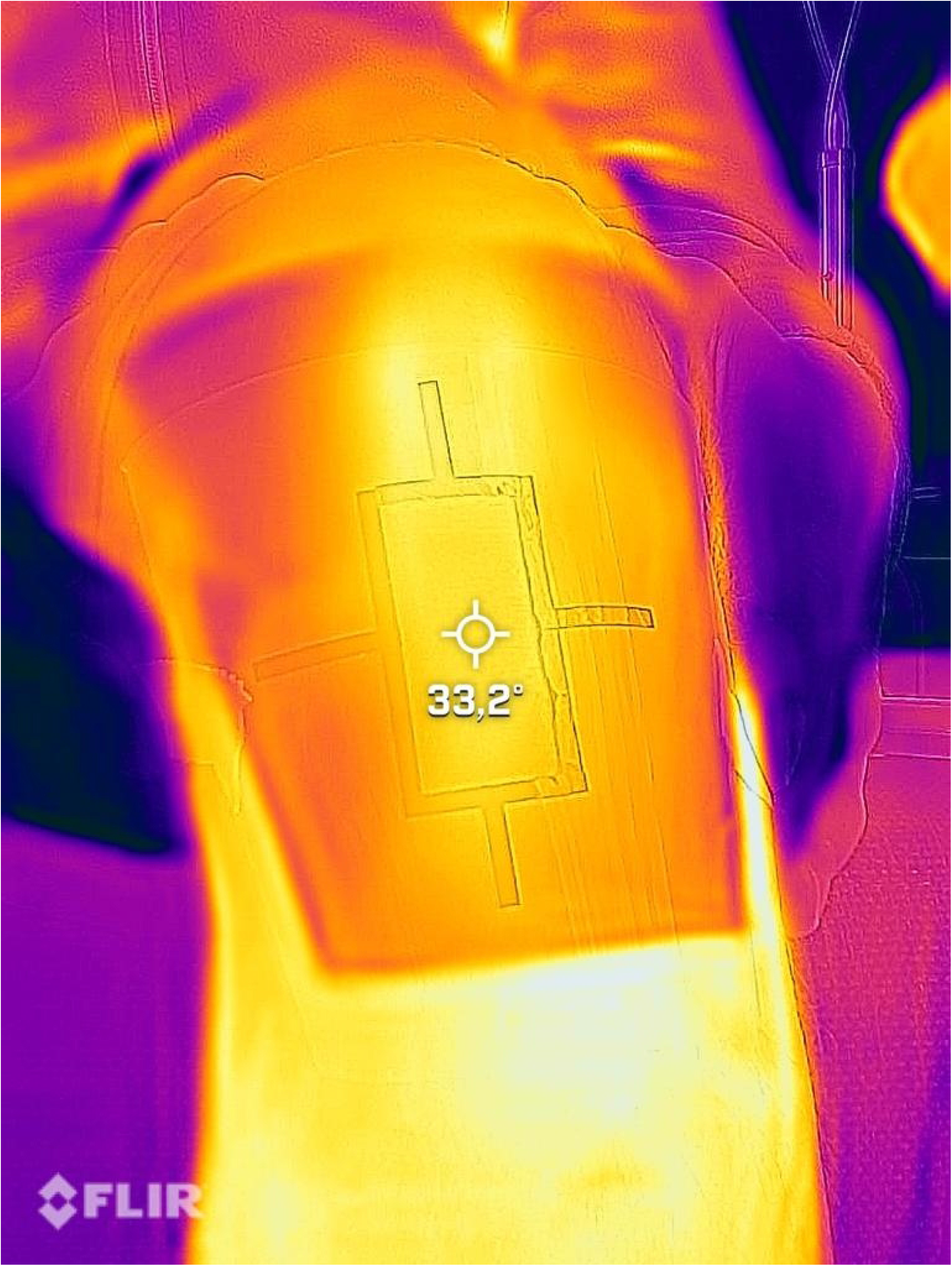
Changes between baseline (t_0_), after treatment (t_1_), and 10 minutes later (t_2_). AU, arbitrary units; SO_2_, oxygen saturation; rHb, relative hemoglobin concentration. Error bars show the standard error of the mean value. Significant at the level * < 0.05; *** < 0.001; **** < 0.0001.

## Discussion

To the authors’ knowledge, this is the first study to investigate the influence of HILT on patellar tendon microcirculation. The results here showed significant changes in all measured parameters.

The thermal changes, which were recorded by infrared thermography, were as expected. There was an increase of 9.45° Celsius immediately after HILT application and a reduction to 1.94° Celsius at follow-up compared to the baseline measurement.

Blood flow increased both immediately after intervention and at follow-up (86.38 AU and 25.76 AU, respectively). These values not only showed statistical significance, they also exceeded the minimum detectable changes of 9.2 AU determined by Wezenbeek et al. [3], although the authors determined these values on the Achilles tendon and the transferability to the patellar tendon is debatable. Furthermore, an increase in SO_2_ was also observable, which was 20.14% after the intervention and 13.48% at follow-up. This improvement of the capillary blood supply could follow a vasodilatation by thermal influences [18]. However, this should also have caused an increase in rHb. In a study preceeding this one, which investigated the influences of purely local thermal changes on the patellar tendon on the microcirculation, an increase in rHb was observed [17]. Nevertheless, the results of this study show a different picture, which is why HILT-specific mechanisms of influence can also be discussed here.

In laboratory studies, there is some evidence of changes in erythrocytes and platelets due to laser therapy that significantly affect flow characteristics. Thereby, the blood viscosity decreases and certain stress adaptation of erythrocytes occurs [9–11]. The phenomenon of an apparent decrease in relative hemoglobin observed in the study would also be consistent with this. The rHb value was decreased by 6.67 AU after the intervention and by 7.9 AU at follow-up. To the authors’ knowledge, this response has not been observed in other studies and may also indicate improved microcirculatory flow properties that increase blood flow velocity in the venous-capillary vasculature. This would explain that the O2C BF meter detects a faster clearance of erythrocytes, leading to a decrease in rHb [19]. However, it should be critically noted in these considerations that although statistically significant results are available here, the observed value changes are relatively small, and it seems questionable whether the minimum detectable changes were exceeded.

Recent studies have also identified red cell changes in long-COVID-19 patients that could affect microcirculation and lead to typical long-COVID-19 symptoms such as fatigue and muscle weakness [20,21]. Therefore, in addition to the scattered work on laser therapy as a method to treat long-COVID-19, work focusing on HILT-specific enhancement of erythrocyte flow properties is of great interest here [12,13].

The study had the strengths mentioned above, but some limitations should be taken into account. For example, it was conducted in a quasi-experimental design that was unblinded and not placebo-controlled. The study group was relatively young, physically active, and of normal weight. Therefore, transferability of the study results to other populations is limited. The results should be understood under the premise of a basic scientific study.

Significant changes in blood flow could be detected after HILT application, which also exceeded the minimal detectable changes. Here, HILT may have an effect beyond mere vasodilation, improving the flow properties of erythrocytes and platelets. Further research investigating these relationships and a controlled design with a comparison group receiving heat-only application represent promising future study approaches.

## Conclusion

This is, to the authors’ knowledge, the first study to evaluate the effect of HILT on the microcirculation of the patellar tendon. It found a significant improvement in blood flow and oxygen saturation after HILT intervention, which were still detectable 10 minutes later. On the other hand, the relative amount of hemoglobin decreased significantly, which could indicate an acceleration of blood flow due to improved flow properties of erythrocytes and platelets.

## Data Availability

All relevant data are within the manuscript and its Supporting Information files.

## Supporting information

S1 Data.

(XLSX)

